# Feasibility of non-invasive nitric oxide inhalation in acute hypoxemic respiratory failure: potential role during the COVID-19 pandemic

**DOI:** 10.1101/2020.05.17.20082123

**Authors:** Kiran Shekar, Sneha Varkey, George Cornmell, Leanne Parsons, Maneesha Tol, Matthew Siuba, Mahesh Ramanan

## Abstract

Acute hypoxemic respiratory failure (ARF) is characterized by both lower arterial oxygen and carbon dioxide tensions in the blood. First line treatment for ARF includes oxygen therapy – intially admininstered non invasively using nasal prongs, high flow nasal cannulae or masks. Invasive mechancial ventilation (IMV) is usually reserved for patients who are unable to maintain their airway, those with worsening hypoxemia, or those who develop respiratory muscle fatigue and consequent hypercapnia.

Inhaled nitric oxide (iNO) gas is known to improve oxygenation in patients with ARF by manipulating ventilation-perfusion matching. Addition of iNO may potentially alleviate the need for IMV in selected patients. This article demonstrates the feasibility of this technique based on our experience of patients with hypoxemic ARF. This technique may also be considered for patients with hypoxic ARF in setting of COVID-19.

## Introduction

Acute respiratory failure (ARF) is a common and life-threatening consequence of a diverse group of diseases ^1,2^. When ARF patients fail conventional oxygen therapies (COT) using non invasive delivery systems (nasal prongs, cannulae, masks) invasive mechanical ventilation (IMV) is often initiated. As new diseases emerge and novel therapies are developed, IMV is becoming more commonplace across the world and its use is rapidly increasing over time^3,4^. This can have an impact on patient outcomes, notwithstanding the burden posed on critical resources. Mechanically ventilated patients represent approximately 3% of acute hospitalisations and 30% of intensive care unit (ICU) admissions ^1,2,5,6^. In the USA, the mean hospital length of stay for a ventilated patient is 14 days with a total hospital cost of $34,257 ^5^. This amounts to $27.0 billion or 12% of all hospital costs for 700,000 patients receiving IMV annually ^2^. Outcomes following IMV are highly dependent on the aetiology of the ARF, severity of illness, plus patient age and co-morbidities. 30% to 40% of patients requiring IMV die without ever being discharged from hospital^1,2^, and many survive with a significantly compromised quality of life ^7-9^. This is most pronounced in the older population - patients over 65 years of age receive IMV at rates that are 3 to 5 times the national average ^2^ with a 2-year survival as low as 48% in the very elderly. Reducing the risks and costs of critical care, of IMV in particular, is a major priority for care providers, health system administrators, tax payers and policymakers^5^.

Avoiding IMV also obviates need for intravenous sedation and neuromuscular blocking agents (NMBA). In addition, IMV is associated with costly complications such as ventilator-associated pneumonia, critical illness weakness, sinusitis, and line sepsis^5^. Avoiding these consequences may reduce the length of ICU stay, complications of prolonged hospitalisation, and subsequently reduce healthcare expenditure. To this end, techniques such as delivering humidified oxygen air mixture at high flow (HFO2) through a nasal cannula^10^ and non-invasive positive pressure ventilation (NIV) have been investigated as potential alternatives to IMV. A recent randomised clinical trial ^11^ in patients with non-hypercapnic, hypoxemic ARF, demonstrated a non significant reduction in the need for IMV when HFO2 therapy was used first, compared with COT and NIV. The use of HFO2 also resulted in an increased 90-day survival. In this study, the rates of subsequent intubation for HFO2, COT and NIV treated patients were 38%, 47% and 50% respectively. The leading cause of intubation in all three groups was worsening ARF and hypoxemia (> 70%).

Hypoxemia in some of these patients can be corrected, at least in part, by the addition of NO gas to the nasal HFO2. Inhaled NO augments oxygenation by improving ventilation-perfusion matching^12^ and, in addition, reduces pulmonary vascular resistance, thereby improving right ventricular performance. Significant improvements in oxygenation have been reported in infants with ARF on nasal continuous positive airway pressure and NO gas inhalation^13^. This study reported that safe ambient nitrogen dioxide (NO2) and NO levels were observed (0.30 and 0.01 ppm). The authors concluded that non-invasively inhaled NO may have a synergistic effect in conjunction with airway recruitment strategies such as nasal CPAP. Inhaled NO does not reduce systemic vasuclar resistance or significantly affect systemic blood pressures-important as many patients with ARF may also present with a sepsis related vasoplegic syndrome.

The use of inhaled NO is of potential relevance in the treatment of patients who develop ARF due to infection with the novel SARS-CoV-2 virus, as the pandemic has led to an unprecedented surge in hospital and intensive care admisisons around the world ^14-16^. Pneumonia is a common presentation in COVID-19 infected patients, with hypoxemic ARF being the most common indication for hospitalisation and subsequent ICU admisison^17-20^. Global numbers indicate that approximately 14% of patients develop severe illness requiring oxygen therapy and 5% will require ICU management. An ICU admission rate of 16 % has been reported from Italy, with a majority of these patients receiving IMV.^14^ Once the disease has progressed to requiring ICU care and mechanical ventilation mortality is significant - 65-76%, and 97% in the over 65 age group^15,16,21-25^.

## Case Series

In this article we present our preliminary experience of using HFO2+NO therapy in patients with hypoxic ARF in a tertiary cardiothroracic ICU between October 2015-July 2019. Data was collected retrospectively, ethics approval was obtained from the local institutional ethics committee (LNR/2019/QPCH/52375) and patient consent was not required. In this period, a total of 197 patients with an admission diagnosis of pneumonia received HFO2 therapy. At clinicians discrerion, 19 patients received HFO2+NO therapy during this period. Patients treated with NO inhalation had a partial pressure of arterial oxygen (PaO2) to fraction of inspired oxygen (FiO2) ratio of < 200 mm Hg on HFO2 therapy. They were able to maintain their airway and clear airway secretions, were haemodynamically stable or receiving low dose vasoactive support. Patients who demonstrated signs of respiratory fatigue were excluded. A summary of patient demographics, severity of illness scores, details of respiratory support provided and outcomes can be found in Table 1. Patients received antibiotics where appropriate, restrictive fluid therapy, nutrition, general supportive care and physical rehabilitation as able.

**Table 1.** Patient demography, details of respiratory support provided and outcomes

|  | HFNC + iNO | HFNC + iNO + subsequent IMV |
| --- | --- | --- |
| **Demographics and physiology** |  |  |
| n | 14 | 5 |
| Males n(%) | 11 (76%) | 2 (40%) |
| Age [median, (IQR)] | 58 (55-70) | 62 (40-70) |
| APACHE-2 Score [median, (IQR)] | 21 (17-24) | 18 (12-24) |
| ANZROD | 0.18 (0.09-0.25) | 0.15 (0.09-0.61) |
| PaO<sub>2</sub>:FiO<sub>2</sub> ratio* (pre- iNO) | 124 (101-154) | 145 (111-187) |
| PaO<sub>2</sub>:FiO<sub>2</sub> ratio* (post<sup>§</sup>- iNO) | 167 (116-230) | 143 (121-224) |
| Respiratory rate b/min (pre-iNO) | 33 (26-36) | 30 (26-33) |
| Respiratory rate b/min (post<sup>§</sup>-iNO) | 22 (19-29) | 22 (17-27) |
| PaCO<sub>2</sub> (mmHg, pre-iNO) | 32 (29-40) | 33 (28-53) |
| PaCO<sub>2</sub> (mmHg, post<sup>§</sup>-iNO) | 33 (31-39) | 33 (26-45) |
| Haemoglobin (g/L) | 105 (93-141) | 132 (117-142) |
| White cell count (x10<sup>9</sup>/L) | 12.7 (6.5-18.4) | 16.8 (8.3-22.9) |
| Creatinine (micromol/L) | 122 (102-145) | 191 (88-303) |
| Urea (mmol/L) | 12.3 (6.5-18.4) | 11.3 (5.4-13.3) |
| Bilirubin (micromol/L) | 17 (10-26) | 20 (14-49) |
| **Diagnosis** |  |  |
| Viral pneumonitis | 5 (36%) | 0 |
| Bacterial pneumonia | 4 (29%) | 2 (40%)<sub>2</sub> |
| Non-cardiogenic pulmonary oedema | 4 (29%) | 3 (40%) |
| Parasitic pneumonia | 1 (7%) | 0 |
| **Treatment** |  |  |
| iNO duration hrs [median, (IQR)] | 81 (25-125) | 203 (51-401) |
| IMV duration hrs [median, (IQR)] | 0 | 172 (148-214) |
| HFNC hrs duration [median, (IQR)] | 89 (40-156) | 413 (202-592) |
| VA ECMO n(%) | 0 | 1 (20%) |
| VV ECMO n(%) | 0 | 1 (20%) |
| **Outcomes** |  |  |
| ICU LOS hrs [median, (IQR)] | 549 (338-1174) | 585 (290-789) |
| Hospital LOS hrs [median, (IQR)] | 734 (505-1267) | 848 (597-1235) |
| Survival to hospital discharge<sup>#</sup> n(%) | 12 (86%) | 5 (100%) |
| Discharged home n(%) | 8 (57%) | 3 (60%) |
| Discharged to other hospital n(%) | 2 (14%) | 0 |
| Discharged to rehabilitation or chronic care facility n(%) | 2 (14%) | 2 (40%) |

APACHE - Acute Physiologic Assessment and Chronic Health Evaluation ; ANZROD- The Australian and New Zealand Risk of Death ; $\text{PaO}_2:\text{FiO}_2$ ratio - ratio of arterial partial pressure of oxygen to fraction of inspired oxygen ; iNO- inhaled nitric oxide; IMV- Invasive mechanical ventilation; HFNC- high flow nasal cannula; VA ECMO- Venoarterial extracorporeal membrane oxygenation ; VV ECMO- Venovenous extracorporeal membrane oxygenation; LOS- Length of stay. \* prior to commencement of inhaled nitric oxide; # two patients who died were considered not suitable for invasive mechanical ventilation. § 4 hours post NO inhalation

Five out of 19 (26%) patients required IMV after a trial of HFO2+NO. The 14 (75%) patients who succesfully avoided IMV after HFO2+NO therapy had slightly higher illness severity as measured by APACHE-2 score (21 vs 18) and lower PaO_2_:FiO_2_ ratio prior to initiation of NO. There was an incement seen in PaO_2_:FiO_2_ ratio following NO inhalation and a decrement in respiratory rate was also noted. Overall survival amongst the HFO2+NO group was 86%, with 2 patients dying who were considered not suitable candidates for IMV. Both these patients maintained autonomy, were able to participate in decision making and spend time with their families prior to initiation of comfort oriented care. Patients who received HFO2+NO only, avoiding IMV, had a shorter ICU and thus hospital length of stay. The patients who eventually received IMV spent signficantly longer time on HFO2+NO therapy prior to intubation, however all survived. Our work unit guideline with regards to HFO2+NO setup and device details can be found in supplement 1. Methaemoglobin levels in blood were monitored regularly and staff reporetd no adverse affects of NO exposure when caring for these patients.

## Discussion

Non-invasive NO inhalation appears to be a feasible alternative to IMV in selected patients with *de novo* hypoxemic ARF. Here, a high flow nasal cannula was used to deliver the gas mixture, however NO can be also delivered via facemasks or through a NIV mask in hypoxic patients. NO use was at the clinicians discretion and therefore further studies are indicated to explore the efficacy and economic analysis of non-invasive NO inhalation in pateints with hypoxic ARF. Whilst studies of the use of NO in acute respiratory distress syndrome (ARDS) have so far failed to demonstrate a survival benefit, the benefits and risks of non-invasive NO inhalation in self-breathing ARF patients has not been systematically studied. Further studies that demonstrate and evaluate the efficacy of less-invasive therapies to improve oxygenation in ARF are warranted. Increased usage of such therapies will have potential impacts on patient outcomes, hospital resources and health economic aspects. There are onging studies evaluating the efficacy of non-invasive NO inhalation to prevent or shorten duration of IMV in COVID-19 patients^26^ as well as those exploring potential viricidal effects of NO in COVID-19 ^27^ During the 2002-2003 SARS epidemic small scale studies demonstrated a reduced duration of ventilatory support in patients infected with the similar coronavirus who received inhaled nitric oxide^28^; most patients included received non-invasive NO therapy.

For a novel technique to be a feasible, safety of both patient and staff is paramount. Training of nursing and respiratory care staff to safely manage this therapy, as well as immediate availability of medical staff to escalate care, are essential to ensure patient safety. It is important that ambient nitrogen dioxide and NO levels are monitored and maintained within safe limits. A major barrier precluding the increased use NO appears to be the delivery cost. However, an electrical generator that generates NO from air has been developed recently which may make inhaled NO more affordable and accessible, especially in under resourced settings where cylinder NO gas is not easily available^29^. Such developing technologies may have a significant impact on the feasibility and financial benefit of non-invasive NO use in a variety of patient population.

Non-invasive NO inhalation may have a potential role in the current pandemic. The mechanisms behind the ARF seen in COVID-19 are not entirely clear but hypoxemia is a predominant feature of the illness. It is postulated that this is potentially due to a dysregulation of lung perfusion and loss of hypoxic vasoconstriction, in addition to a signficantly increased intrapulmonary shunt fraction. Pulmonary endothelieal involement^30^ and both macro and mcirovasuclar thromboembolic phenomena^31,32^ have also been described. In addition, ARDS patients who are self-breathing there are concerns regarding patient-self inflicted lung injuiry (P-SILI) in the setting of a high respiratory drive in setting of hypoxia.^33^ Inhaled NO may be an useful adjunct to improve ventilation:perfusion matching in this setting to reduce the of degree hypoxemia which may also blunt the respiratory drive. The theoretical risks of P-SILI whilst spontaneously breathing has to be balanced against the known risks of intubation and IMV, including diagphragmatic myotrauma,^34^ sedation and NMBA use, and critical illness weakness. This remains an important area for future research.

In this setting, optimal respiratory support strategies for COVID-19 respiratory failure are yet to be defined. Disappointing survival rates have been reported in patients with increasing age and comorbidites who are intubated for COVID-19 associated ARF^15,16,22,35^. WHO interim guidelines and the Australia and New Zealand Intensive Care Society guidelines recommend the use of HFO2 therapy, with optimal airborne precautions, as a potential strategy to avoid intubation^36,37^. In a recent study, there was no variation in aerosol production in spontaneously breathing volunteers amongst room air, 6 L/min nasal cannula, 15 L/min via non-rebreather mask, 30 L/min HFNC, and 60 L/min HFNC, regardless of coughing^23^. Non invasive NO inhalation along with HFO2 support as described in our patients may be of potential benefit. Equally interesting will be the the effects of prone positioning in self-breathing patients^38^ receiving HFO2+NO therapy.

The outbreak is challenging healthcare systems around the world. In a pandemic, ICU resources are stretched and strategies that can avoid or delay ICU admission are important – both for patients outcomes and to conserve healthcare resources. Treatment options than can prevent or delay disease progression are highly desirable. Oxygen delivery through HFO2 via nasal cannulae or NIV can be done in non-ICU settings to potentially avoid the need for IMV^10^. In a similar fashion, non-invasive NO inhalation may have a potential role during the early phases of COVID-19 infection to prevent progression of the disease to the point of requiring IMV. Additionally amongst patients who are intubated and are demonstrated to be iNO-responsive, earlier extubation may be facilitated using non invasive iNO – thus reducing the duration of IMV and liberating patients from ventilators faster. This would potentially shorten the duration of ICU stay and have important implications on healthcare resources.

## Data Availability

No external datasets provided

## Acknowledgements

Kiran Shekar acknowledges research support from the Metro North Hospital and Health service and the Prince Charles Hospital Foundation. We sincerely thank the nursing staff at the Adult Intensive Care Services, the Prince Charles Hospital and the multidisiciplinary team members involved in care of the patients.

## Financial/nonfinancial disclosures

None declared

**Figure 1.**
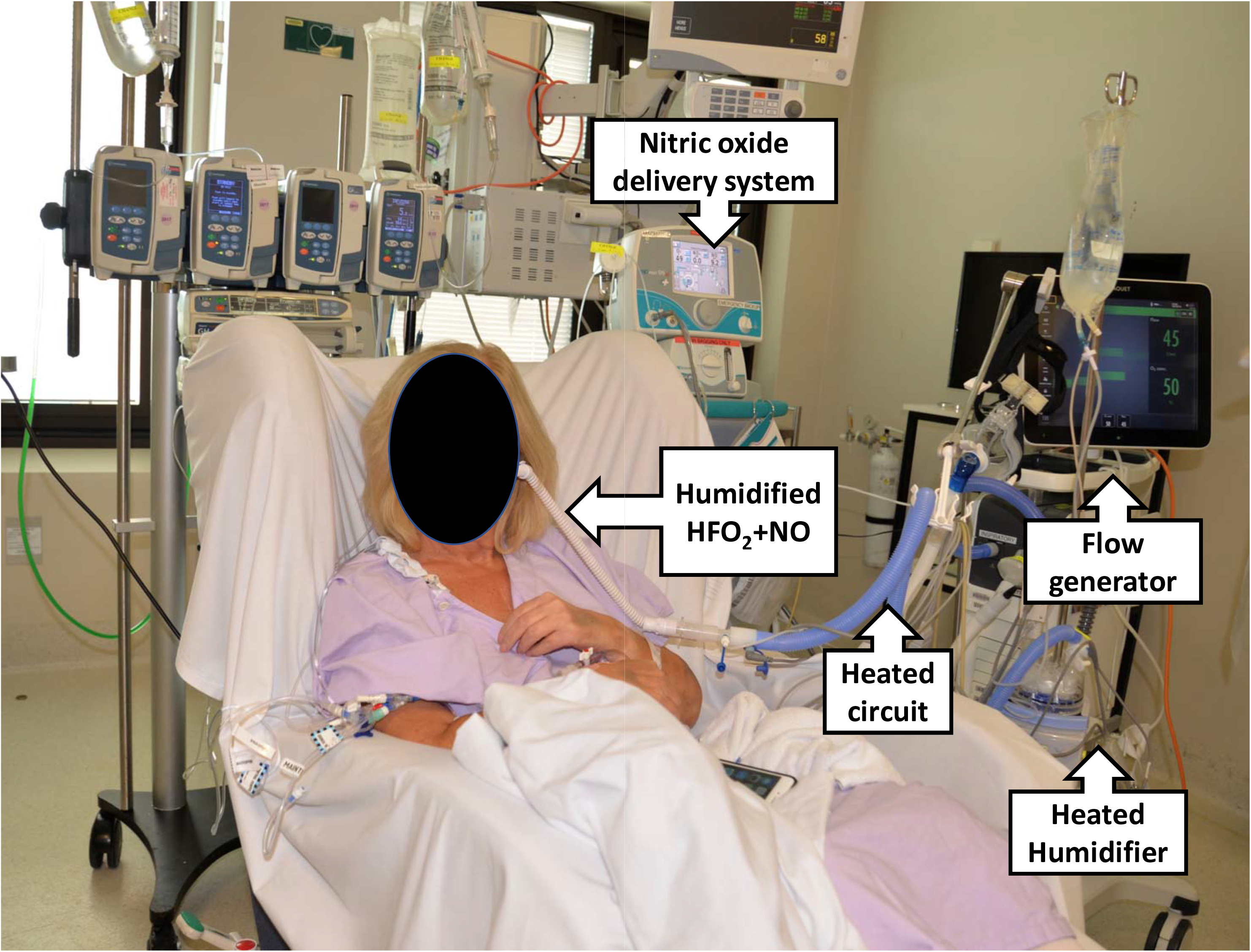
Typical patient supported with humidified high flow air, oxygen and nitric oxide mixture gas mixture delivered via a nasal cannula.

Supplement 1: High flow nasal cannula and nitric oxide set up

